# Association of musculoskeletal pain with incidence and recurrence of cardiovascular disease: a longitudinal cohort in China

**DOI:** 10.1101/2025.06.24.25330239

**Authors:** Yu Chen, Haiqiang Guo, Le Yan, Chuanquan Peng, Yan Liu, Yan Deng, Min Zhu

## Abstract

**Aims:** To examine the association of musculoskeletal pain with both incident and recurrent cardiovascular disease (CVD) in a prospective cohort.

**Methods:** A prospective cohort analysis was performed using a nationally representative dataset of middle-aged and older population in China during 2011-2020. In the final analysis, 8,716 participants were evaluated for incident CVD, and 1,151 were examined for recurrent CVD events. The associations between musculoskeletal pain and both incident and recurrent CVD were examined using Cox proportional hazards regression and restricted cubic spline models.

**Results:** Participants with musculoskeletal pain showed a significantly higher CVD incidence rate than pain-free individuals throughout follow-up (3.45 vs. 2.43 per 100 person-years). Individuals with musculoskeletal pain had a 54% increased risk of recurrence (HR = 1.54, 95% CI: 1.28-1.85). Participants with multisite pain demonstrated progressively greater CVD risks, with significant trend for both incident and recurrent events (P trend <0.001). All pain dynamics (generated, disappeared, intermittent, persistent) were significantly associated with both incident and recurrent CVD. Subgroup analyses revealed elevated risks of both incident and recurrent CVD among females, individuals with high BMI, and urban residents. Participants with hypertension or diabetes showed significantly higher risks of incident CVD.

**Conclusions:** Musculoskeletal pain is independently associated with both incident and recurrent CVD; prolonged pain duration and an increasing number of pain sites further amplify these risks. Musculoskeletal pain should be recognized not merely as a comorbidity but as a modifiable risk factor for cardiovascular health.

## Introduction

Globally, cardiovascular disease (CVD) maintains its position as the foremost cause of death and disability, with incidence rates showing a steady upward trajectory Despite significant advances in prevention and treatment, complex cardiovascular events, with both new-onset and, especially, recurrent cases, continue to pose a major challenge to public health. Approximately 20–30% of patients experience a second event within five years of the initial diagnosis, and these recurrent events are directly associated with increased mortality rates.^1^ ^2^ Emerging evidence suggests that non-traditional modifiable factors, including musculoskeletal pain, may correlate with CVD.

Musculoskeletal pain, a common condition affecting nearly 30% of the global population, is increasingly recognized as a potential contributor to CVD. Prior studies have indicated that musculoskeletal pain is associated with a higher prevalence of CVD and an elevated risk of cardiovascular mortality.^5–9^ However, existing research has several limitations. Most studies have focused on single pain types or specific anatomical regions.^10^ ^11^ Causal inferences cannot be drawn from cross-sectional studies,^12–14^ and many studies are conducted exclusively in developed countries and regions^11^ ^12^ ^13^ ^15^. Furthermore, current research has not fully explored how pain characteristics, such as persistence, intermittency, and resolution, as well as pain distribution (location and multiplicity), influence CVD. Notably, the association between musculoskeletal pain and cardiovascular event recurrence remains underexplored. Given that over 75% of the global CVD burden is concentrated in low-income and middle-income countries and regions, with the heaviest burden in South and East Asia. ^16–18^ Clarifying the associations between musculoskeletal pain and both the onset and recurrence of CVD is critical for developing targeted interventions to reduce the cardiovascular burden in these regions.

This study examines the association between musculoskeletal pain and both incident and recurrent CVD events across a decade of follow-up. Specifically, we examine the relationship between different types of pain dynamics (newly generated, disappeared, intermittent, persistent), pain locations (spine, upper limbs, lower limbs), and multisite pain involvement with both the onset and recurrence of CVD risk.

## METHODS

### Study participants

The present study utilized data from the China Health and Retirement Longitudinal Study (CHARLS), an ongoing nationally representative prospective cohort. Details of CHARLS have been provided elsewhere^19^. In brief, CHARLS is a longitudinal study focused on health, economic and social aspect of adults aged 45 years and older. Initiated in 2011, CHARLS is conducted biennially. We selected 2011 survey as baseline, with follow-up assessment in 2013, 2015, 2018 and 2020. In this study, we excluded participants with missing data of musculoskeletal pain and CVD and those aged <45 years old. We also excluded participants who lost to follow-up. Ultimately, this study included 8716 and 1151 participants in final analysis of incident CVD and recurrent CVD, respectively. Fig. 1 illustrated how participants were included and excluded. Ethical approval for CHARLS was obtained from the Biomedical Ethics Committee of Peking University (IRB0000001052-11015). All participant provided written informed consent.

**Fig. 1.**
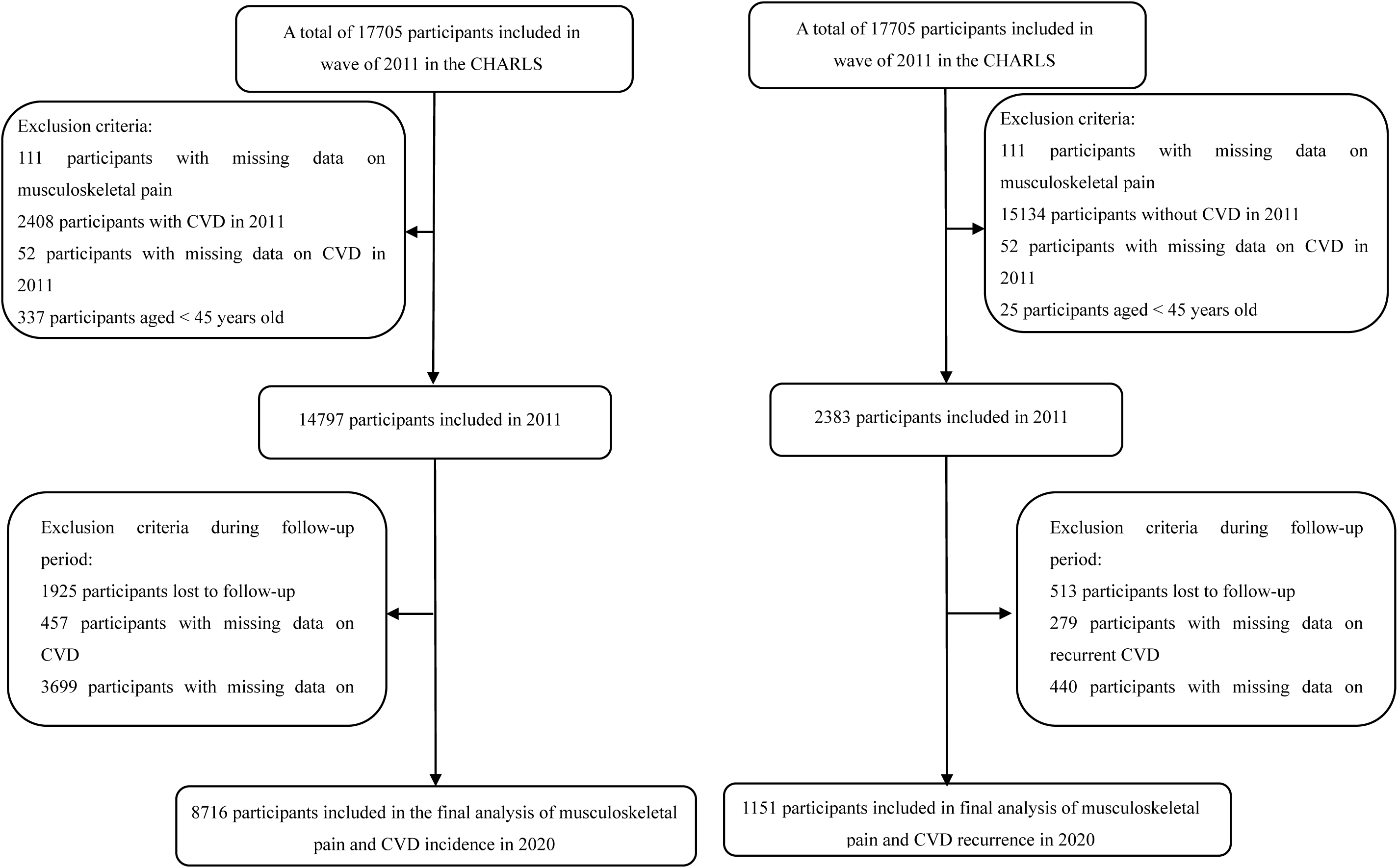
Flow chart of the participants inclusion and exclusion process. CHARLS, China Health and Retirement Longitudinal Study. CVD, cardiovascular disease.

### Assessment of musculoskeletal pain

Musculoskeletal pain was assessed using the CHARLS questionnaires, in which participants were asked to list all areas of the body where they currently experienced pain. Details are provided in Supplementary Table S1. Musculoskeletal pain was categorized into spine pain (neck pain, back pain and waist pain), lower limbs pain (leg pain and knee pain) and upper limbs pain (shoulder pain, arm pain and wrist pain). If participants experienced any of the following: spine pain, lower limbs pain and upper limbs pain, they were classified as having musculoskeletal pain. If participants did not report pain at baseline but developed pain after a follow-up survey, they were categorized as having newly generated pain. Conversely, if participants reported pain but remained pain-free after a follow-up survey, they were classified as having pain that disappeared. Intermittent pain was defined as pain that recurred and disappeared during the follow-up. Persistent pain during follow-up was defined as continuous pain. The total duration of pain was determined by summing the duration of pain across different periods during the follow-up. For individuals who experienced pain consistently throughout all waves, the total duration was calculated from the start to the end of the follow-up period. Conversely, for participants who did not experience pain at any wave, the total duration was designated as 0 years. For participants whose pain appeared or disappeared during follow-up, the total pain duration was calculated as the half of the time between two survey waves.

### Assessment of incidence and recurrence of CVD

Data on participants’ incidence and recurrence of CVD were collected by trained researchers through in-person interviews. CVD status was confirmed using following question at baseline and follow-up period: “Have you been diagnosed with heart attack, coronary heart disease, angina, congestive heart failure, other heart problems, or stroke?” Recurrence of CVD was defined based on the following question during follow-up period: “Have a doctor told you that you had another stroke or a heart attack since last survey?” If participants died, the incidence or recurrence of CVD was determined based on information obtained from the participant’s family, with deaths due to CVD being categorized as CVD incidence or recurrence events.

### Covariates

Following previous literature, potential confounding variables included age (continuous variable), sex (women or men), BMI (continuous variable), residence (urban region or rural region), education level (below primary school or primary school or above), marital status (married status or other status), smoking status (yes or no), drinking status (yes or no), annual household income (<10,000 yuan, 10,001-50,000 yuan, or >50,000 yuan), hypertension (yes or no), diabetes (yes or no), solid fuel use (yes or no), cognitive function score (continuous variable). Solid fuel use was was determined by whether participants used coal, crop residue or wood burning for cooking and heating.

### Statistical analyses

The distributions of participant characteristics were summarized as mean ± standard deviation (SD) for continuous variables and as frequency (percentage) for categorical variables, respectively. The ANOVA or Chi-square tests were used to compare baseline characteristics between the pain and no-pain groups. Cox proportional hazards regression models were used to explore associations between musculoskeletal pain and CVD, as well as recurrence of CVD. To account for potential confounders, we constructed three models: (1) Model 1: unadjusted; (2) Model 2: adjusted for age, sex and BMI; (3) Model 3: Model 2 plus residence, educational level, marital status, smoking status, drinking status, annual household income, hypertension, diabetes, solid fuel use and cognitive function score. Additionally, the restricted spline models with 3 knots were used to evaluate associations between duration of musculoskeletal pain and CVD, as well as its recurrence. The restricted spline models utilized the hazard ratios (HRs) derived from Model 3. Interaction and subgroup analyses were also conducted to explore the relationships between baseline musculoskeletal pain and participants’ characteristics (sex, age, BMI, residence, education, smoking status, drink status, hypertension, and diabetes) using Cox proportional hazard regression models.

We conducted several sensitivity analyses to assess the robustness of our results. First, we used multiple imputation (MI) method to evaluate the effect of missing covariates data. Second, we excluded participants with incident CVD and recurrent CVD within 2 years of follow-up, as these may have been influenced by other risk factors. Third, we excluded participants with a severe cognitive impairment whose cognitive function score below 3 points (larger score indicate better cognitive function, up to a maximum of 21) to address potential recall bias. All data were analyzed using R software (version 4.2.2).

## Results

### Participants characteristics

Participant characteristics at baseline are shown in Table 1, categorized according to the presence or absence of musculoskeletal pain. The final analysis included 8,716 participants for incident CVD and 1,151 participants for recurrent CVD. For incident CVD analysis, the average age of participants was 57.76 years (SD = 8.57). Compared to pain-free participants, those with musculoskeletal pain were significantly older, more likely to be female, reside in rural areas, and had lower BMI, education levels, household income, and cognitive scores. They also showed lower rates of alcohol consumption and smoking, but higher prevalence of hypertension and diabetes, along with greater solid fuel use; In the recurrent CVD analysis, participants averaged 60.33 years (SD = 8.85), with 709 females. Participants without musculoskeletal pain tended to be male, slightly older, urban residents, with higher education attainment, greater alcohol consumption, reduced solid fuel use, and better cognitive performance.

**Table 1.** Participants’ characteristics according to musculoskeletal pain at baseline

| Variable | Cardiovascular disease |  |  |  | Recurrence of cardiovascular disease |  |  |  |
| --- | --- | --- | --- | --- | --- | --- | --- | --- |
|  | Overall | Pain | No pain | *P* value | Overall | Pain | No pain | *P* value |
| N | 8716 | 2452 | 6264 |  | 1151 | 521 | 630 |  |
| Age (years) | 57.76 ± 8.57 | 58.43 ± 8.47 | 57.49 ± 8.60 | <0.001 | 60.33 ± 8.85 | 59.09 ± 8.32 | 61.35 ± 9.15 | <0.001 |
| Sex |  |  |  | <0.001 |  |  |  | <0.001 |
| Women | 4546 (52.2) | 1533 (62.5) | 3013 (48.1) |  | 709 (61.6) | 362 (69.5) | 347 (55.1) |  |
| Men | 4170 (47.8) | 919 (37.5) | 3251 (51.9) |  | 442 (38.4) | 159 (30.5) | 283 (44.9) |  |
| BMI (kg/m<sup>2</sup>) | 23.46 ± 3.55 | 23.33 ± 3.71 | 23.51 ± 3.49 | 0.037 | 24.64 ± 4.08 | 24.71 ± 4.18 | 24.58 ± 4.00 | 0.598 |
| Residence |  |  |  | <0.001 |  |  |  | <0.001 |
| Urban region | 1401 (16.1) | 246 (10.0) | 1155 (18.4) |  | 311 (27.0) | 93 (17.9) | 218 (34.6) |  |
| Rural region | 7315 (83.9) | 2206 (90.0) | 5109 (81.6) |  | 840 (73.0) | 428 (82.1) | 412 (65.4) |  |
| Education level |  |  |  | <0.001 |  |  |  | <0.001 |
| Below primary school | 2380 (27.2) | 839 (34.2) | 1541 (24.5) |  | 317 (27.5) | 174 (33.4) | 143 (22.7) |  |
| Primary school or above | 6336 (72.8) | 1613 (65.8) | 4723 (75.5) |  | 834 (72.5) | 347 (66.6) | 487 (77.3) |  |
| Marital status |  |  |  | 0.131 |  |  |  | 0.588 |
| Married status | 7346 (84.3) | 2043 (83.3) | 5303 (84.8) |  | 959 (83.3) | 438 (84.1) | 521 (82.7) |  |
| Other status | 1370 (15.7) | 409 (16.7) | 961 (15.2) |  | 192 (16.7) | 83 (15.9) | 109 (17.3) |  |
| Smoking status |  |  |  | <0.001 |  |  |  | 0.234 |
| Yes | 3385 (38.8) | 821 (33.5) | 2564 (41.0) |  | 387 (34.4) | 165 (32.6) | 222 (35.9) |  |
| No | 5331 (61.2) | 1631 (66.5) | 3700 (59.0) |  | 764 (65.6) | 356 (67.4) | 408 (64.1) |  |
| Drinking status |  |  |  | <0.001 |  |  |  | 0.033 |
| Yes | 3031 (34.8) | 739 (30.1) | 2292 (36.6) |  | 262 (22.8) | 103 (19.8) | 159 (25.2) |  |
| No | 5685 (65.2) | 1713 (69.9) | 3972 (63.4) |  | 889 (77.2) | 418 (80.2) | 471 (74.8) |  |
|  | Overall | Pain | No pain | *P* value | Overall | Pain | No pain | *P* value |
| Annual household income (yuan) |  |  |  | 0.002 |  |  |  | 0.403 |
| <10000 | 8127 (93.2) | 2324 (94.8) | 5803 (92.6) |  | 1105 (96.1) | 501 (96.2) | 604 (95.9) |  |
| 10001-50000 | 524 (6.0) | 113 (4.6) | 411 (6.6) |  | 42 (3.6) | 17 (3.2) | 25 (3.9) |  |
| >50000 | 65 (0.7) | 15 (0.6) | 50 (0.8) |  | 4 (0.3) | 3 (0.6) | 1 (0.2) |  |
| Hypertension |  |  |  | <0.001 |  |  |  | 1.000 |
| Yes | 1648 (19.0) | 550 (22.6) | 1098 (17.6) |  | 569 (49.6) | 257 (49.6) | 312 (49.6) |  |
| No | 7068 (81.0) | 1902 (77.4) | 5166 (82.4) |  | 582 (50.4) | 264 (50.4) | 318 (50.4) |  |
| Diabetes |  |  |  | 0.007 |  |  |  | 1.000 |
| Yes | 370 (4.3) | 127 (5.2) | 243 (3.9) |  | 123 (10.8) | 56 (10.8) | 67 (10.7) |  |
| No | 8346 (95.7) | 2325 (94.8) | 6021 (96.1) |  | 1028 (89.2) | 465 (89.2) | 563 (89.3) |  |
| Solid fuel use |  |  |  | <0.001 |  |  |  | <0.001 |
| Yes | 6185 (71.6) | 1970 (81.2) | 4215 (67.9) |  | 822 (72.9) | 407 (79.6) | 415 (67.3) |  |
| No | 2531 (28.4) | 482 (18.8) | 2049 (32.1) |  | 329 (27.1) | 114 (20.4) | 215 (32.7) |  |
| Cognitive function score | 10.60 ± 4.31 | 9.49 ± 4.20 | 11.04 ± 4.27 | <0.001 | 10.61 ± 4.34 | 9.80 ± 4.37 | 11.31 ± 4.20 | <0.001 |

Statistical significance $p < 0.05$

### Longitudinal association between musculoskeletal pain and both incident CVD and recurrent CVD

Table 2 presented the associations between musculoskeletal pain and risks of incident and recurrent CVD. During follow-up, participants with musculoskeletal pain showed a significantly higher incidence rate of CVD than those without (3.45 per 100 person-years vs. 2.43 per 100 person-years). After full adjustment for potential confounders (Model 3), musculoskeletal pain correlated with a 37% greater likelihood of developing CVD (HR=1.37, 95% CI: 1.24-1.52). Significant associations were consistently observed across all pain sites. We found evidence of a graded relationship between pain site multiplicity and CVD risk (*P* trend <0.001), and participants with 5-6 pain sites had the highest risk of new-onset CVD (HR=1.55, 95% CI: 1.25-1.91).

**Table 2.**
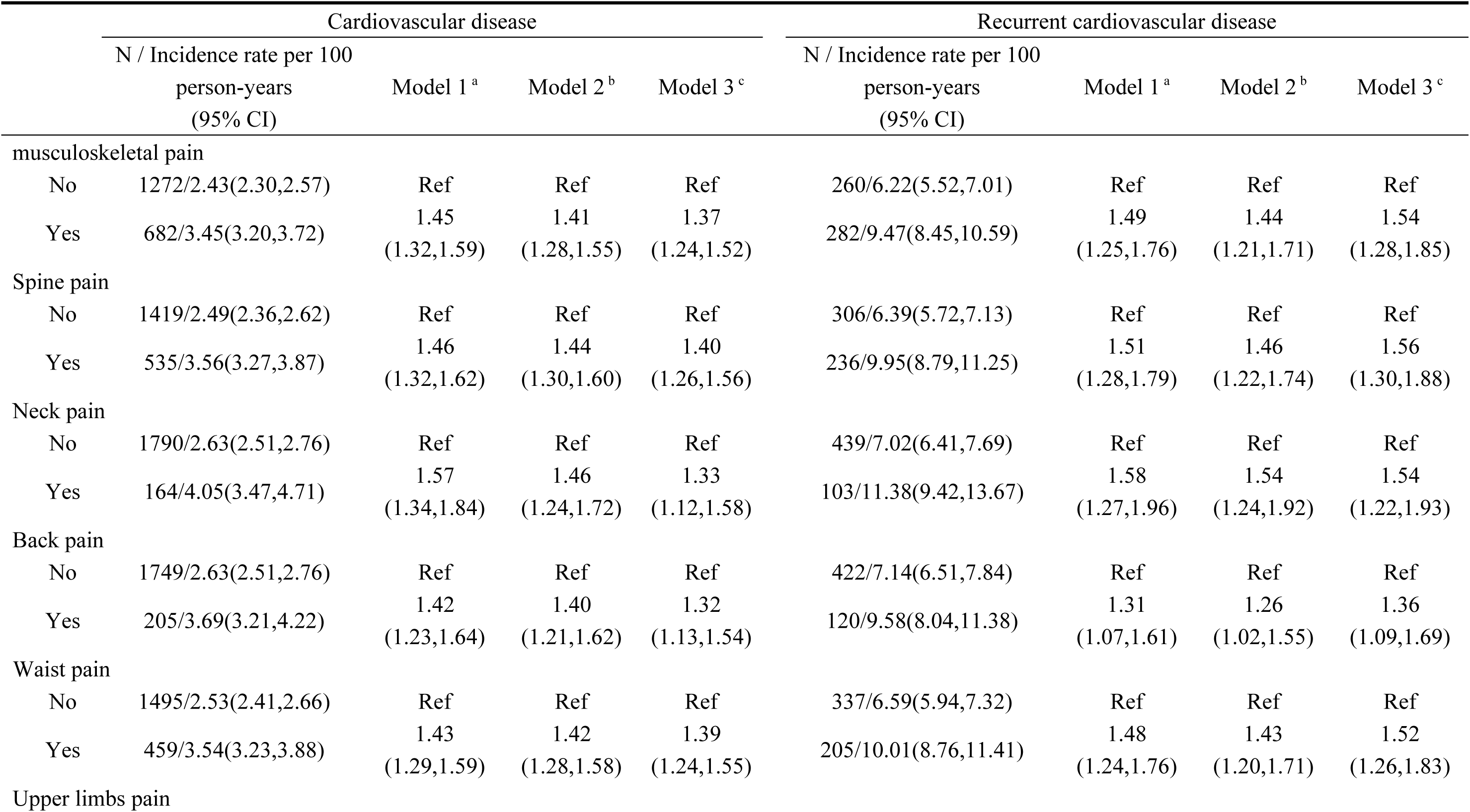
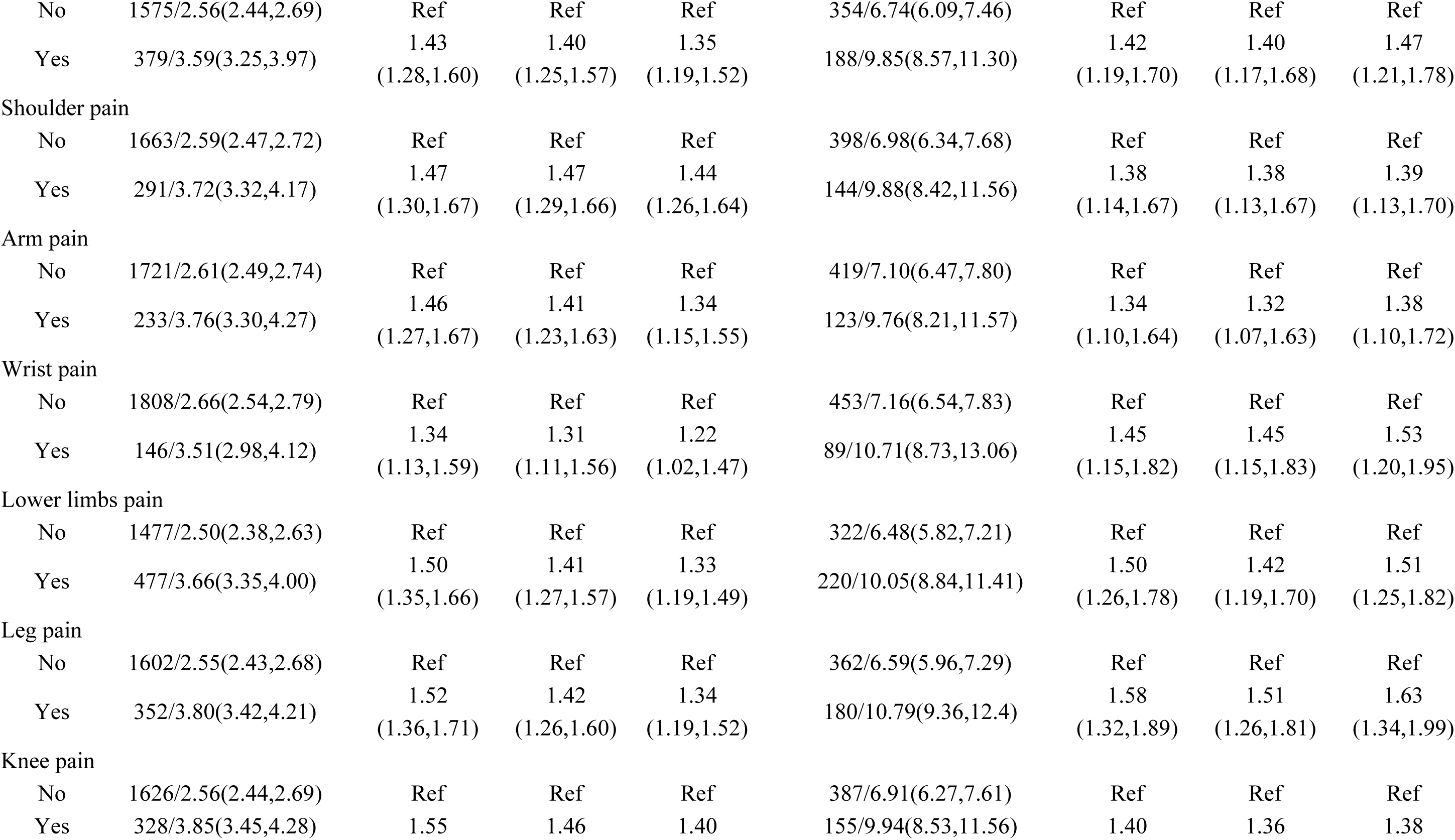
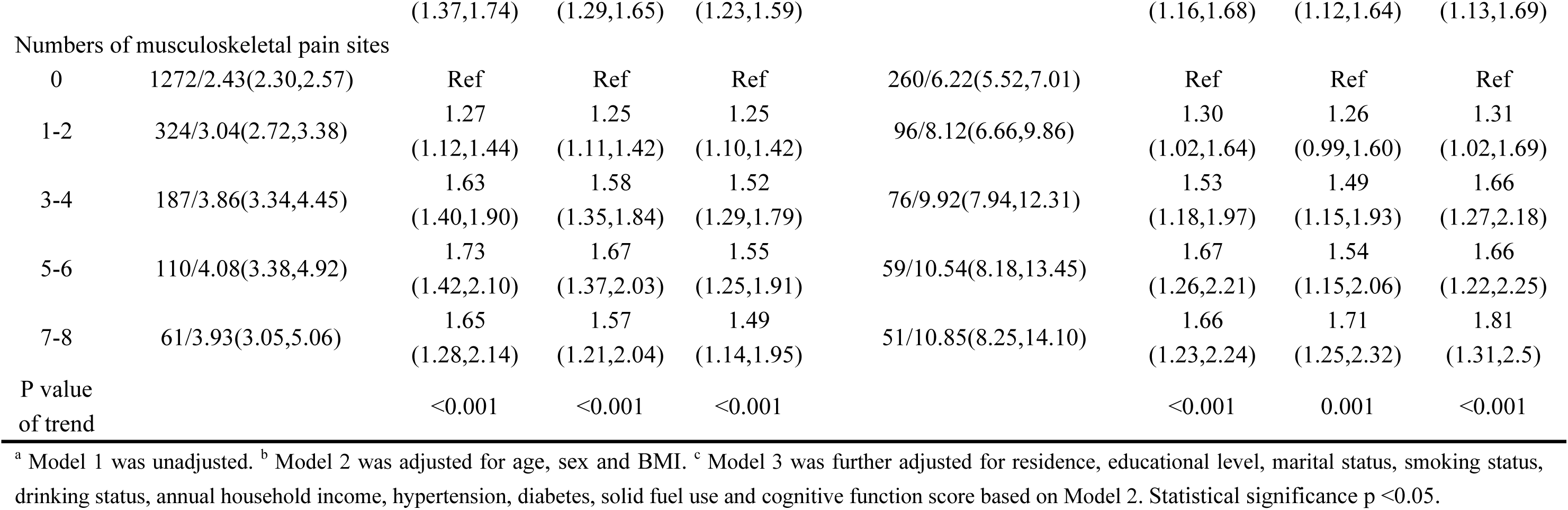
Associations of musculoskeletal pain at baseline with both incident and recurrent cardiovascular disease

Similarly for recurrent CVD, participants with musculoskeletal pain demonstrated elevated recurrence rates. After full adjustment, musculoskeletal pain independently predicted 54% increased recurrence risk (HR=1.54, 95% CI: 1.28-1.85). A significant exposure-response relationship was observed, with recurrence risk tending to increase as the number of pain sites increased. (*P* trend <0.001). Compared to pain-free participants, adjusted HRs were 1.31 (95% CI: 1.02-1.69) for 1-2 sites, 1.66 (95% CI: 1.27-2.18) for 3-4 sites, 1.66 (95% CI: 1.22-2.25) for 5-6 sites, and 1.81 (95% CI: 1.31-2.50) for 7-8 sites.

### Association between pain dynamics and incident CVD and its recurrence

As detailed in Table 3, distinct pain dynamics exhibited graded associations with cardiovascular outcomes. All pain dynamics (generated, disappeared, intermittent, persistent) were significantly associated with both incident and recurrent CVD. Notably, persistent pain demonstrated the strongest effect, with HR of 2.68 (95% CI: 2.19-3.27) for incident CVD and 3.23 (95% CI: 2.19-4.77) for recurrent CVD. Participants whose pain had disappeared showed increased risks of both incident CVD (42% higher risk) and recurrent CVD (88% higher risk) compared to those without pain history. Among all pain sites, persistent neck pain exhibited particularly high hazard ratios, indicating a strong link with cardiovascular events.

**Table 3.**
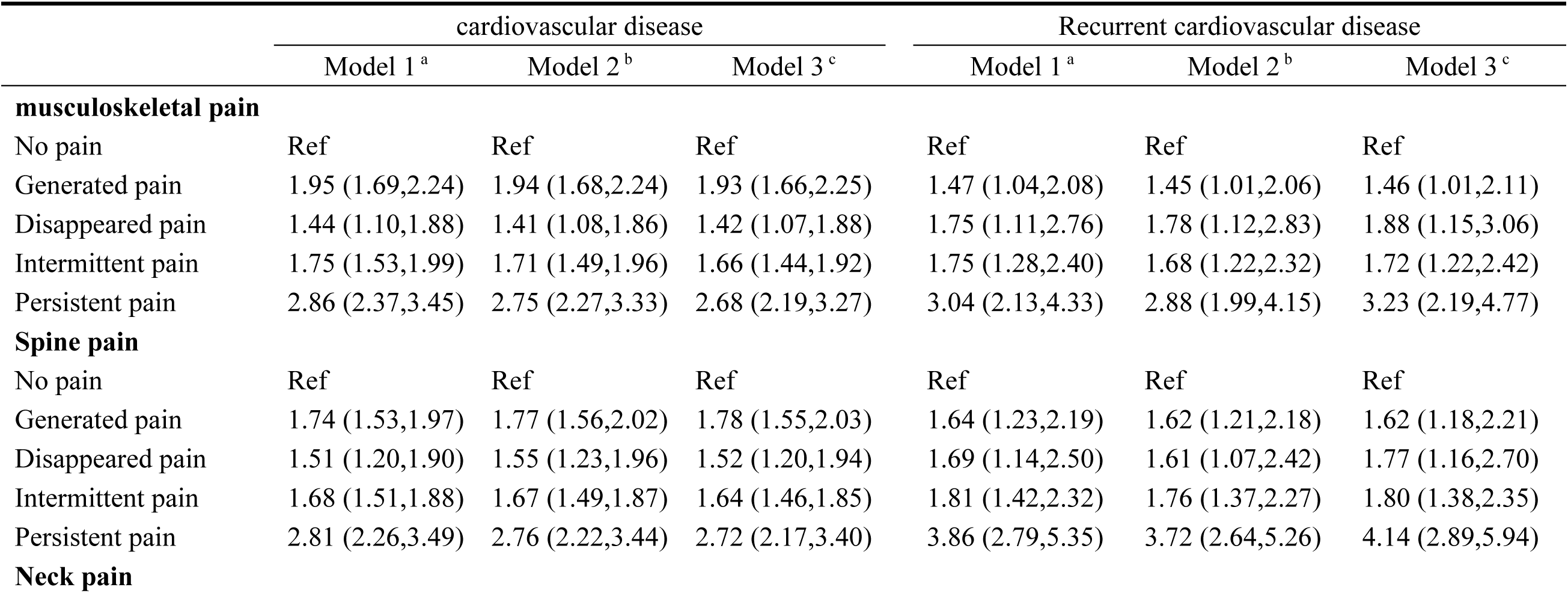
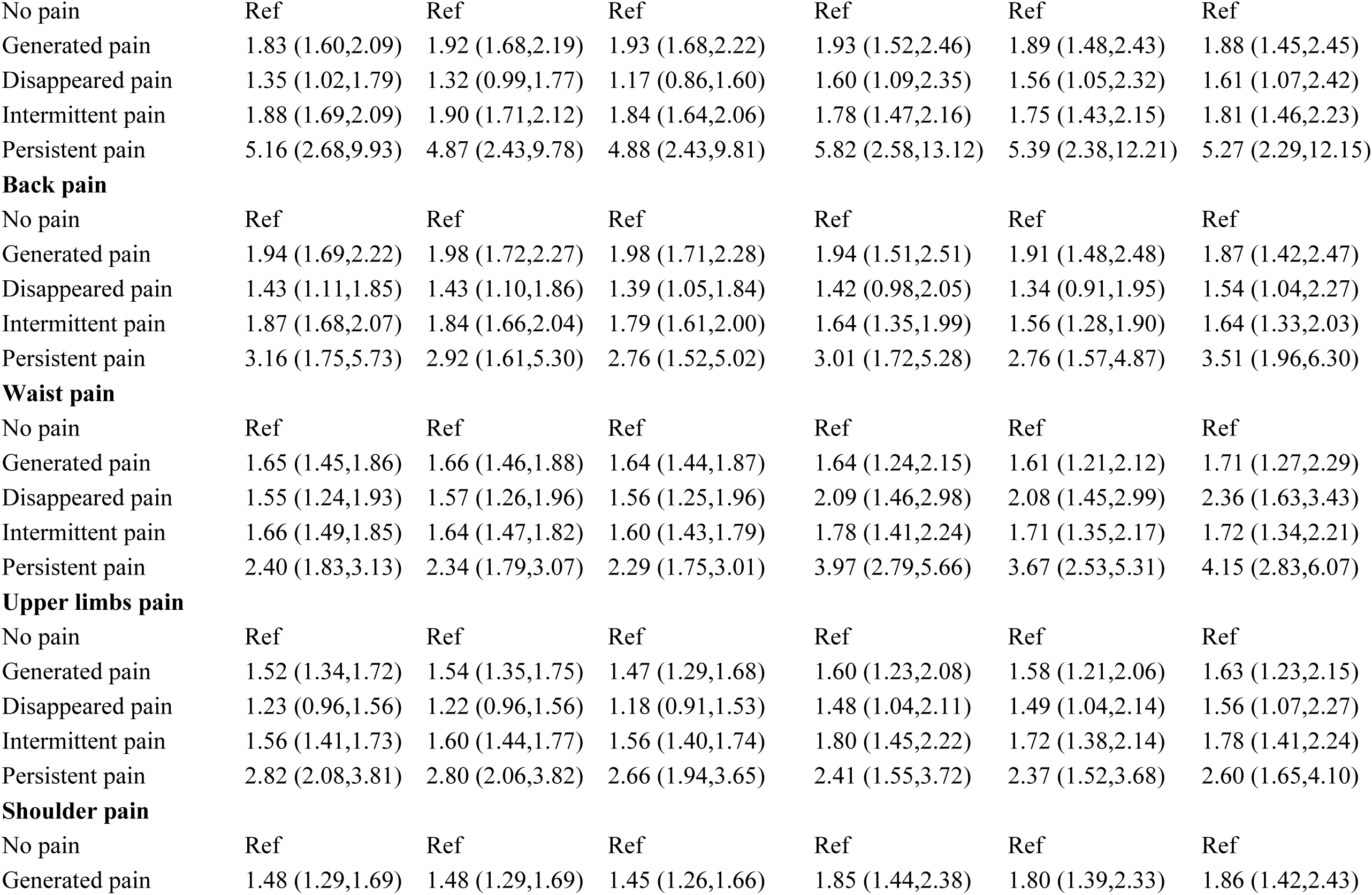
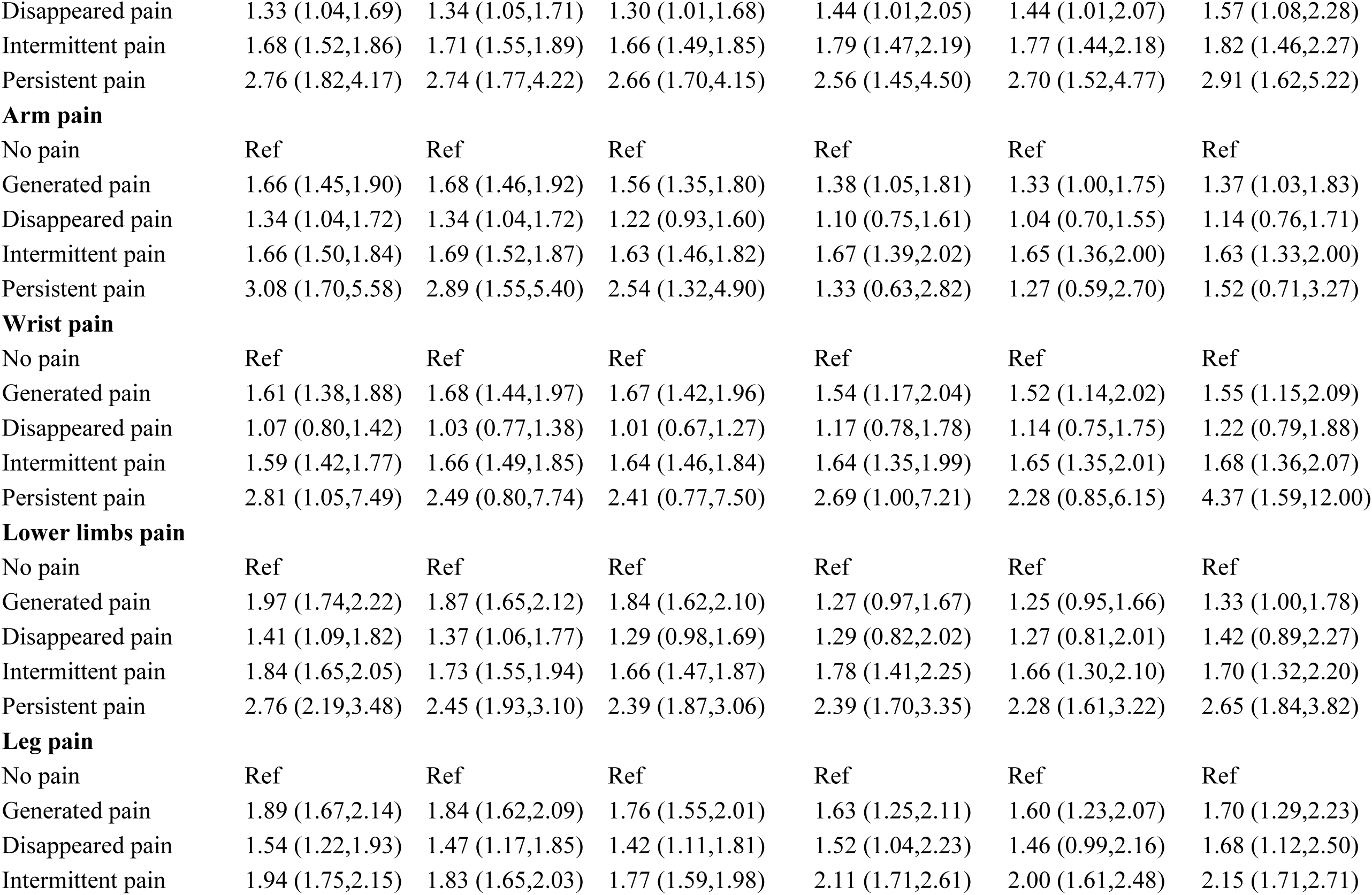
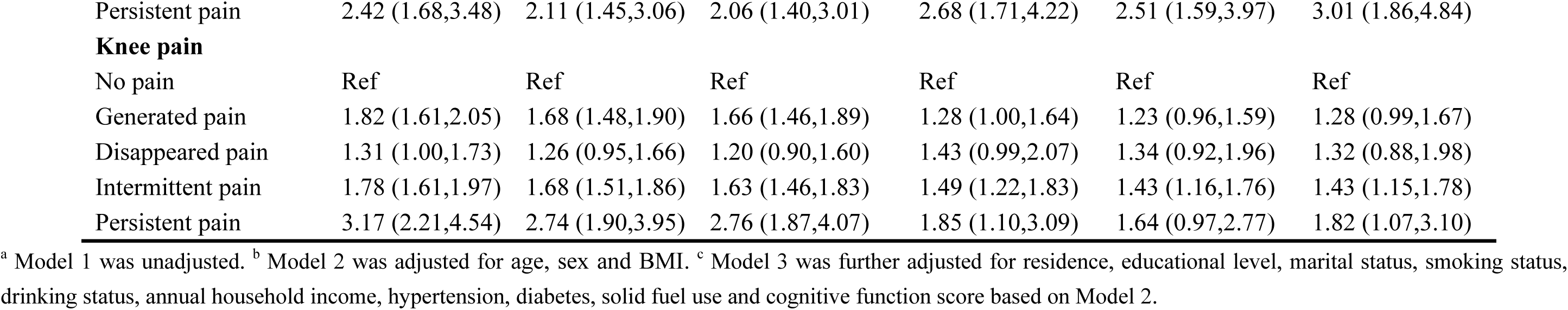
Associations between musculoskeletal pain dynamics during follow-up and both incident and recurrent cardiovascular disease

Figures 2 and 3 display the restricted cubic spline analyses examining the relationships between musculoskeletal pain duration and incident/recurrent CVD, respectively. We observed linear associations between pain duration and incident CVD for most sites (musculoskeletal overall, lower/upper limbs, spine, shoulders, knees, arms, waist, and wrists), while leg and back pain durations showed nonlinear relationships (*P* < 0.05). For recurrent CVD, pain duration showed linear associations across all specific anatomical sites.

**Fig. 2.**
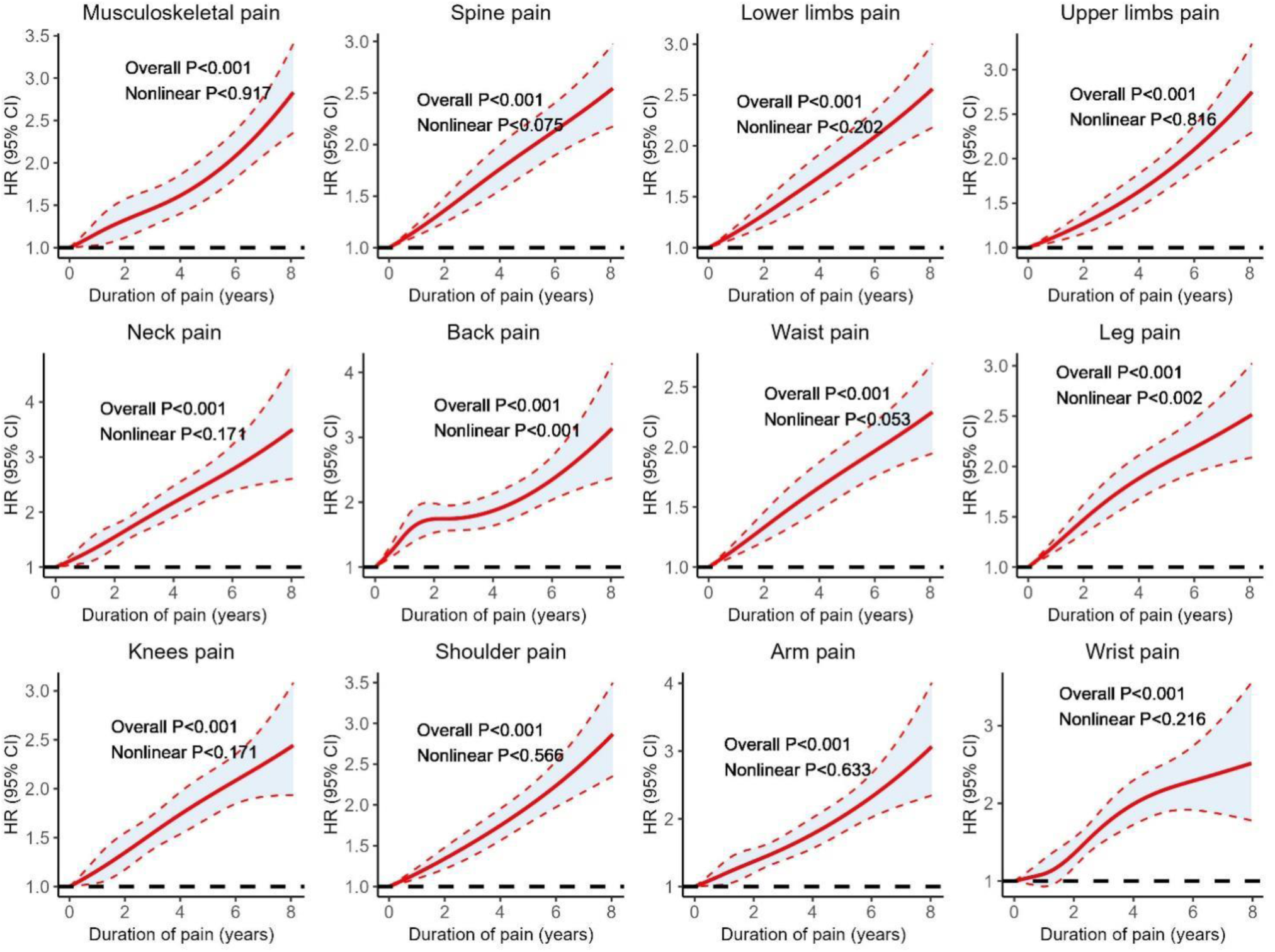
Associations between musculoskeletal pain duration and incident cardiovascular disease

**Fig. 3.**
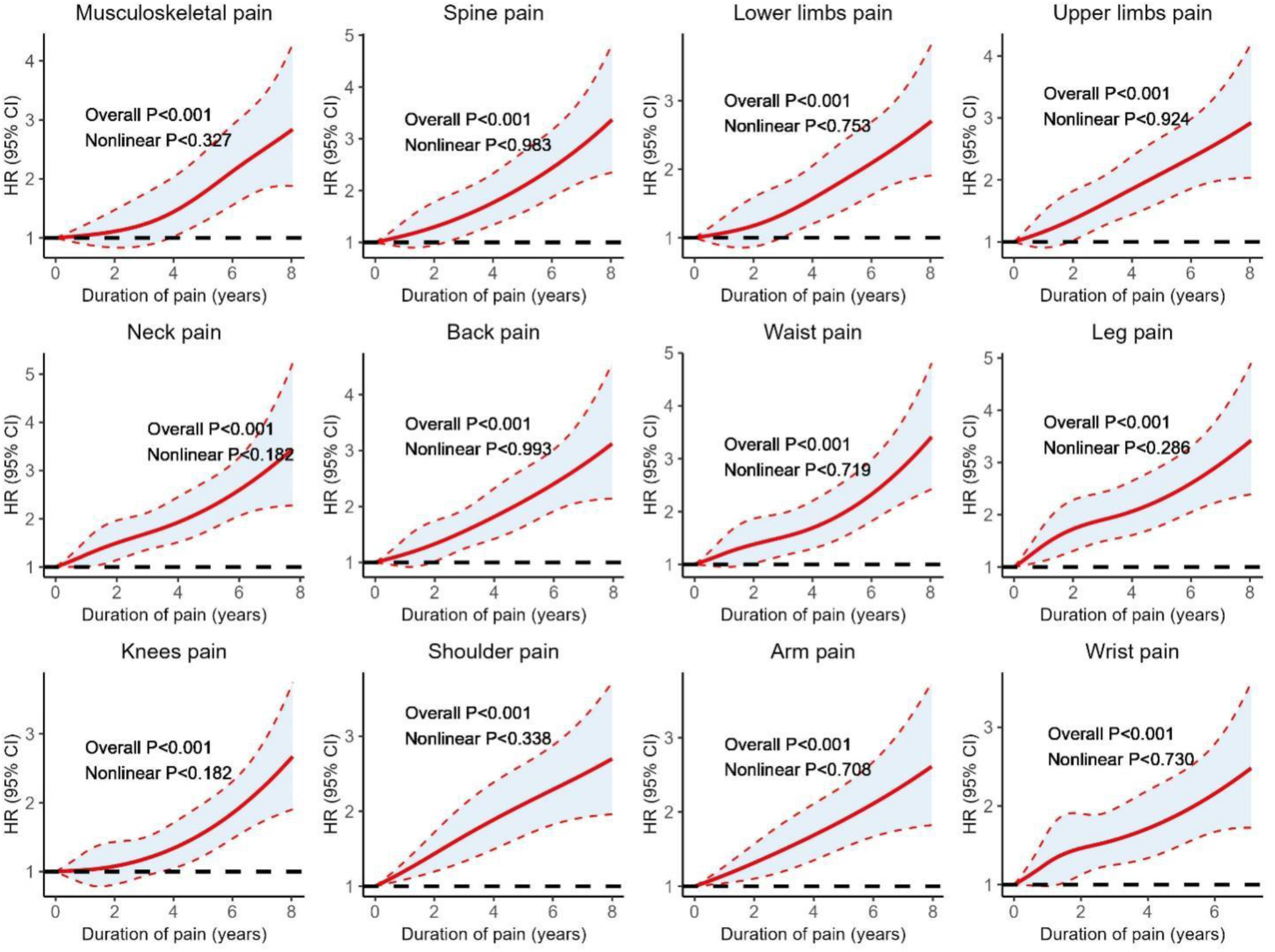
Associations between musculoskeletal pain duration and recurrent cardiovascular disease

### Interaction and subgroup analyses

Figure 4 presents the interaction and subgroup analyses examining the associations between musculoskeletal pain and participant characteristics for both incident and recurrent CVD. No statistically significant interaction effects were found between musculoskeletal pain and any participant characteristics. However, Subgroup analyses revealed elevated risks of both incident and recurrent CVD among females, individuals with high BMI, and urban residents. Furthermore, participants with hypertension or diabetes showed significantly higher risks of incident CVD.

**Fig. 4.**
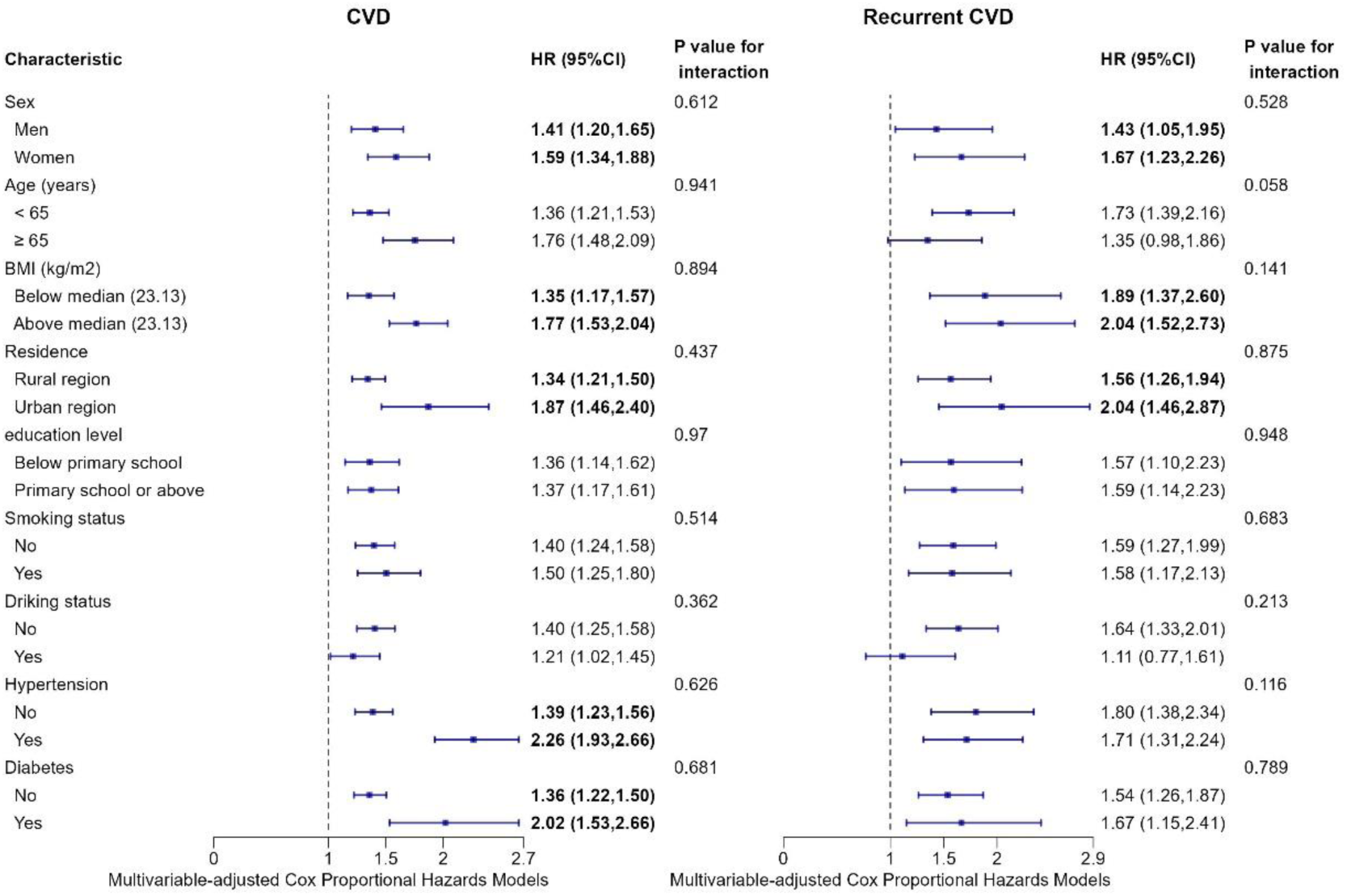
Interaction and subgroup analyses between musculoskeletal pain and participants’ characteristics on both incident and recurrent CVD. CVD, cardiovascular disease. Statistical significance p <0.05.

### Sensitivity analyses

The positive associations between musculoskeletal pain and both incident and recurrent CVD demonstrated consistent robustness across all sensitivity analyses (Tables S2-S4). After applying multiple imputation (Table S2), all musculoskeletal pain sites maintained significant associations with elevated risks of both incident and recurrent CVD (all *P* < 0.05). These associations persisted (Table S3) after excluding participants who developed incident CVD or experienced recurrence within the initial biennial follow-up. Similarly, after excluding participants with cognitive function scores less than 3 points (total cognitive function scores: 21 points), musculoskeletal pain remained significantly associated with both incident and recurrent CVD risks (Table S4).

## Discussion

Leveraging longitudinal data from the CHARLS, this study provides compelling epidemiological evidence linking musculoskeletal pain to elevated risks of both incident and recurrent CVD in a developing country context. After comprehensive adjustment for sociodemographic, lifestyle, and clinical confounders, musculoskeletal pain maintained independent associations with a 37% increased incidence and 54% higher recurrence risk of CVD. These robust associations were consistently observed across all specific anatomical sites, including spinal, lower limb, and upper limb pain. Notably, we identified a potential exposure-response gradient, in which an increase in the number of pain sites and prolonged pain duration were associated with elevated CVD risk. These findings together suggest that musculoskeletal pain exerts a cumulative cardiovascular burden, with both multisite involvement and chronicity amplifying CVD susceptibility.

Our finding that a positive association between musculoskeletal pain and incident CVD has been supported by previous studies. Rönnegård et al. using data from the UK Biobank found that individuals aged 40-69 years suffering from chronic pain had an increasing risk for cardiovascular events with increasing widespreadness^20^. In Taiwanese studies utilizing the National Health Insurance Research Database, fibromyalgia, a chronic and widespread musculoskeletal pain, was linked to a 47% increased risk of coronary heart disease (HR=1.47, 95% CI: 1.43–1.51) and a 25% greater risk of overall stroke (HR=1.25, 95% CI: 1.21–1.30)^22^. Notably, a study by Tsai et al.^10^ reported an even stronger association, with fibromyalgia patients exhibiting a 2.11-fold elevated risk of coronary events (HR=2.11, 95% CI: 1.46–3.05). Additionally, several studies showed that people with pain due to osteoarthritis were at increased risk for CVD^23–26^. In this study, we focused on the pain symptoms rather than underlying pathophysiological mechanisms of the pain. These findings collectively highlight that musculoskeletal pain could impact CVD development.

Our study extended previous findings by demonstrating that an increased number of pain sites was exposure-responsively associated with an elevated probability of incident and recurrent CVD in a Chinese middle-aged and older population. These results aligned with Tian et al. (2023)^27^, who reported that each additional chronic pain site elevated myocardial infarction risk by 12% (HR=1.12 per site) in the UK Biobank cohort, and Tian et al. (2024)^28^, who found that widespread chronic pain directly correlated with increased arterial stiffness (β=0.06 per site) and reduced left ventricular ejection fraction (β=−0.17 per site). Supported by previous research^20^ ^27^ ^28^, our results highlighted that multisite musculoskeletal pain strongly amplifies the risk of CVD.

Our analysis of pain dynamics revealed that prolonged pain duration significantly increased CVD risk, with elevated risk persisting even after pain disapperance. Our findings suggested that persistent pain had the strongest positively association with incidence and recurrence of CVD. Despite the disappearance of pain, affected individuals maintained elevated risks of incident CVD and recurrent CVD, implying that prior musculoskeletal pain may induce lasting cardiovascular alterations independent of current symptoms. Consistent with our results, previous researches have reported chronic pain independently predicted elevated CVD risk.^20^ ^28–30^ Rönnegård et al. found that individuals with pain disapperance (duration of pain not >3 months) had a 13% higher risk of a composite CVD outcome than individuals with no pain. ^20^ Our finding provided evidence that the disappearance and reappearance of pain can both lead to cardiovascular risk, regardless of the duration of pain. Patients with a history of musculoskeletal pain, particularly persistent pain, should be flagged for closer cardiovascular monitoring, as these individuals may benefit from early interventions to mitigate CVD risk.

Remarkably, we examined CVD recurrence in a developing country context. To the best of our knowledge, no previous study had specifically demonstrated the association between musculoskeletal pain and recurrent cardiovascular events. Our findings uniquely highlighted that pain burden (both site number and duration) profoundly predicted CVD recurrence. Recurrence of cardiovascular events significantly increased patient mortality^31–33^, with the risk of death rising significantly as the number of recurrences grew, and each recurrence adding approximately 66.6% to the risk.^31^. Moreover, widespread pain was also correlated with an increased hazard ratio for cardiovascular mortality.^8^ This interaction suggests that pain-driven CVD mortality may operate through recurrent events, as prolonged pain duration may exacerbate vascular dysfunction over time. Crucially, in resource-limited settings where complete pain resolution is often unattainable, our finding that even partial reduction of pain sites lowers recurrence risk offers a pragmatic strategy. This strategy involves targeting high-risk pain patterns, such as persistent pain, which could disrupt the cycle of recurrence-mortality, independent of achieving full pain remission. By bridging the gap between pain management and secondary CVD prevention, our work redefines chronic pain not merely as a comorbidity, but as a modifiable determinant of post-CVD survival.

Several possible mechanisms could underlie the observed association between musculoskeletal pain and CVD. First, chronic pain is associated with heightened sympathetic nervous system activity^29^ ^30^, increased levels of inflammatory markers^34^ ^35^, which have been implicated in atherosclerosis and cardiovascular events. Second, individuals with musculoskeletal pain may adopt sedentary lifestyles due to reduced physical activity, which was a well-established risk factor for CVD (36). Third, the psychological distress associated with chronic pain, including anxiety and depression, may exacerbate cardiovascular risk through a series of complex biological mechanisms, such as increased sympathetic activity, endothelial dysfunction, thrombotic mechanisms, and dysregulation of the hypothalamic–pituitary–adrenal axis.^37^ ^38^ Additionally, the use of certain medications for pain management, such as nonsteroidal anti-inflammatory drugs (NSAIDs), has been linked to adverse cardiovascular outcomes.^39^ ^40^

The strengths of this study lie in its large-scale sample and longitudinal design covering a ten-year period, and comprehensive adjustment for confounding variables. Our study is pioneering in systematically exploring the longitudinal association between musculoskeletal pain and recurrent CVD in developing countries. The repeated measurements of pain and CVD outcomes across multiple waves reduce the likelihood of reverse causality bias compared to cross-sectional designs.

Several limitations should be acknowledged. First, both musculoskeletal pain and CVD status are self-reported, which might lead to potential recall bias or misclassification. However, given that pain is subjective, self-reported pain assessments remain a valuable and widely used method for capturing pain experiences in both research and clinical settings. Second, our findings may not be generalizable beyond the Chinese population aged 45 years and older, and further validation in diverse populations is warranted. The absence of data on detailed medication use and genetic predispositions, has also restricted our ability to examine the relationship between musculoskeletal pain and CVD risk. Third, the study design does not capture precise pain duration measurements, which limites our ability to determine whether pre-disapperance pain duration proportionally influenced post-disapperance CVD risk.

### Conclusions

Musculoskeletal pain is independently associated with both incident and recurrent CVD; prolonged pain duration and an increasing number of pain sites further amplify these risks. Our findings underscore the importance of recognizing musculoskeletal pain not merely as a comorbidity but as a modifiable risk factor for cardiovascular health.

### Author contribution

YC performed data scrub and analysis. MZ wrote the initial draft, and reviewed and edited the manuscript. YD formulated the research goals and aims. YL designed methodology and tested the existing code components. All authors had unrestricted access to data and contributed to the interpretation of results, critical revision of the manuscript and approved the final manuscript.

## Conflict of interest

## Data Availability

Please contact the corresponding Author to obtain all data in the manuscript

